# A simple feed forward neural network to predict the 2025 outbreak of measles in the USA

**DOI:** 10.1101/2025.04.01.25325011

**Authors:** Grace Burtenshaw, Georgina McCallum, Ruethaichanok Kardkasem, Kirsty R. Short, Meagan Carney

## Abstract

**Background:** Measles is a highly contagious viral disease associated with a variety of severe complications. Since 1963, widespread usage of a highly effective vaccine has made measles a largely preventable disease. However, recent rises in vaccine hesitancy in the United States has seen increasing incidence of measles cases, including an ongoing (22^nd^ March 2025) outbreak originating in Gaines County, Texas.

Accurate modelling is essential for informing public the health response to measles outbreaks. Current strategies are limited by a variety of challenges primarily associated with obtaining adequate real-time data in conjunction with an active outbreak.

**Methods:** Here, we use a dynamic time warping (DTW) approach, to facilitate the use of historical outbreak data to model the ongoing measles outbreak in the United States. Data from various historical outbreaks were used as features for two neural network architectures: a feedforward neural network (FNN) and a novel, biologically informed neural network (BINN). The latter BINN architecture incorporated dynamics from populations of susceptible (S), infected (I) or recovered (R) individuals, commonly described in association with SIR infectious disease models.

**Findings:** Both FNN and BINN architectures performed well across 34-weeks of testing data, predicting measles cases with a mean squared error of less than 2 (1.1060 and 1.1451, respectively). Additionally, a 5-week forward prediction of case numbers closely matches CDC estimates, as reported on 22^nd^ of March 2025. Interestingly, no difference in the accuracy of forward predicted case numbers was observed between FNN and BINN architectures.

**Interpretation:** Overall, this study highlights the value of historical data in combination with relatively simple FNN architecture for accurately modelling ongoing or emerging measles outbreaks. Such modelling strategies represent an essential tool for future outbreak management and a timely reminder of the importance of consistent public health responses (vaccination) to otherwise preventable diseases.

**Funding:** KRS is supported by an NHMRC Investigator Grant (2007919)

**Research in context:** We searched for the terms “MACHINE LEARNING” AND “MEASLES” AND “OUTBREAK” in Google Scholar. Numerous articles have reported the use of machine learning for the prediction of measles outbreaks. However, no study to date has used dynamic time warping and historical data for feature selection and training data in order to develop a simple feed-forward neural network.

**Added value of this study:** Here, we develop a new, rapid and highly accurate methodology for predicting measles outbreaks in the USA in real time. We use this model to show that number of future measles cases will likely increase over time, highlighting the importance of continued and consistent management (vaccination) of otherwise preventable diseases.

**Implications:** The present study provides a new approach to infectious disease modelling that circumvents the need for diverse and granular datasets for feature selection. We use this novel approach to show that without a dramatic shift in public health policy and vaccine advocacy the number of measles cases in the USA will continue to rise.

## INTRODUCTION

Measles is a highly contagious viral disease that often results in severe complications such as pneumonia, encephalitis, and even death ^1^. In addition to acute complications, Measles is also associated with numerous long-term sequelae including subacute sclerosing panencephalitis (SSPE), generalised immunosuppression, hearing loss and chronic lung pathologies ^2,3^. The primary component of the public health response to this disease is a highly effective vaccine, licensed for use in the 1960s ^2^. Since the introduction of this intervention, the incidence of measles has declined by >95% ^2^. Accordingly, in the year 2000, measles was declared eliminated in the United States (US), with no evidence of continuous transmission for over 12 months ^4^. Unfortunately, since 2019, the number of reported measles cases in the US has been increasing. This coincides with increasing vaccine hesitancy and political polarisation of attitudes towards vaccination, which has seen first-dose vaccination rates drop as low as 90% in children (<35 months of age)^5^.

At the start of 2025, a new outbreak of measles was recorded in Gaines County, West Texas. This outbreak centred around the under-vaccinated, close-knit local Mennonite community. At the time of writing (March 22^nd^), there have been 378 cases of measles recorded in the US in 2025, as recorded on the Centre for Disease Control (CDC) web portal. This includes approximately 300 cases associated with the Texas outbreak, which has now spread to New Mexico, Pennsylvania, Oklahoma and California and resulted in at least two deaths. Predicting the course of these measles outbreaks is essential to help public health officials allocate resources efficiently, implement targeted vaccination campaigns and prevent further spread.

Traditionally, infectious disease outbreaks are predicted using SIR models, which categorize populations into Susceptible (S), Infected (I), and Recovered (R) groups to simulate disease spread ^6^. These models use mathematical equations to estimate infection rates, peak outbreak times and the effectiveness of public health interventions. More recently, machine learning, rather than SIR models, has been used to predict disease outbreaks due to the capacity for analysis of complex patterns in large, real-world datasets.

There are numerous examples of machine learning being used to inform our understanding of measles outbreaks ^7-9^. However, such studies have sought to use a wide variety of features associated with the demography and geography of an outbreak region (e.g. vaccination coverage, socioeconomic status, international travel volume, exposure history/contact history) to predict real-time outbreak dynamics ^7-9^. In the case of a current, ongoing measles outbreak, this approach limits clinical translatability as granular data collation is not the priority of first responders, thus delaying the generation of a predictive model. Analysis of historical data as machine learning features offers the opportunity to incorporate all these factors without the time-consuming need for current, granular, local data. However, there are numerous limitations associated with using historical data (local or international) to train machine learning models. Firstly, incomplete coverage across selected proposed prediction features results in reductions to usable training and testing cohorts. Solutions to this, such as the generation of synthetic data to address class imbalances pose its own challenges ^7^. More significantly, there are challenges associated with time series matching, where timing differences between historical and current outbreak dynamics may preclude alignment, reducing the size of available training and test sets ^8^.

In the context of typical infectious disease outbreaks various factors can alter the actual or recorded infection dynamics of a pathogen; for example, the emergence of viral mutations, changes in diagnostic criteria or regional differences in public health policies/response. However, such limitations do not necessarily apply to Measles virus ^10^. Here, viral mutations are limited, diagnostic criteria are well established and the public health response (vaccination) to outbreaks has been consistent for decades ^1,4^.

As such, with use of dynamic time warping to address problems of time series matching, a limited set of historical (international) measles outbreak data may be used to rapidly and accurately predict the ongoing Gaines County outbreak with a simple feed-forward neural network. The success of this approach represents an invaluable tool for incorporation into the public health response to current and future measles outbreaks.

## MATERIALS AND METHODS

### Data sources

Monthly reported measles cases from 1998 − 2024 were collected from the European Centre for Disease Prevention and Control (ECDC) on 12 March 2025. Cases from Austria and Belgium were the focus of this study due in part to the similar levels of measles vaccinations in these countries and the USA (Supplementary Figure 1). The Austrian and Belgium timeseries of monthly reported cases were examined for measles outbreak onset and conclusion. A full outbreak was defined as the observation of a small number of measles cases (0-1) resulting in a peak number of measles cases, followed by a decline to a small number of cases (0-1) once again. If cases did not reach near zero on decline, the outbreak was extended to include as many peaks as necessary until a true near zero decline was observed. Twelve historical outbreaks in total were used to form the feature dataset: 5 from Austria (years 2013/14, 2014/15, 2018/19/20, 2022/23, 2023/24) and 7 from Belgium (2003/04, 2007/08, 2011/12, 2014/15, 2015/16, 2016/17, 2017-20, 2023/24). One final feature vector was created using a one-week time delay of the weekly CDC reported measles cases from 1 January 2023 to 1 March 2025 accessed on 6 March 2025.

Total weekly reported measles cases across the United States were obtained through the CDC web portal as a timeseries of observations from 1 January 2023 to 1 March 2025 accessed on 6 March 2025.

### Curating the feature matrix

Given that the historical outbreak timeseries for feature extraction occur on the order of one month, we employed a method called dynamic time warping to align our time series to the reference weekly measles timeseries that we are interested in forecasting. Dynamic time warping works by finding the most optimal alignment of each historical timeseries, or path of points, to our reference timeseries by calculating the total distance between the reference and historical timeseries for every possible alignment in the space. Supplementary Figure 2 illustrates this process for one of our historical monthly outbreak timeseries.

Owing to the high infection rate of measles, all measles outbreak timeseries often follow the trend of runs of low values followed by sudden spikes in cases which makes learning and forecasting the timeseries an almost impossible task from a modelling perspective. Traditional approaches to addressing this issue involve some version of signal smoothing through a windowed average; however, this makes forecasting estimates unreliable for individual outbreaks ^11-13^. A significant benefit to using historical measles outbreaks as feature data is that they provide a change of basis, converting the original highly complex function describing weekly measles cases in the USA against time to a smoother function describing weekly measles cases in the USA against a warped feature timeseries where the network can be easily constructed (Figure 1).

**Figure 1:**
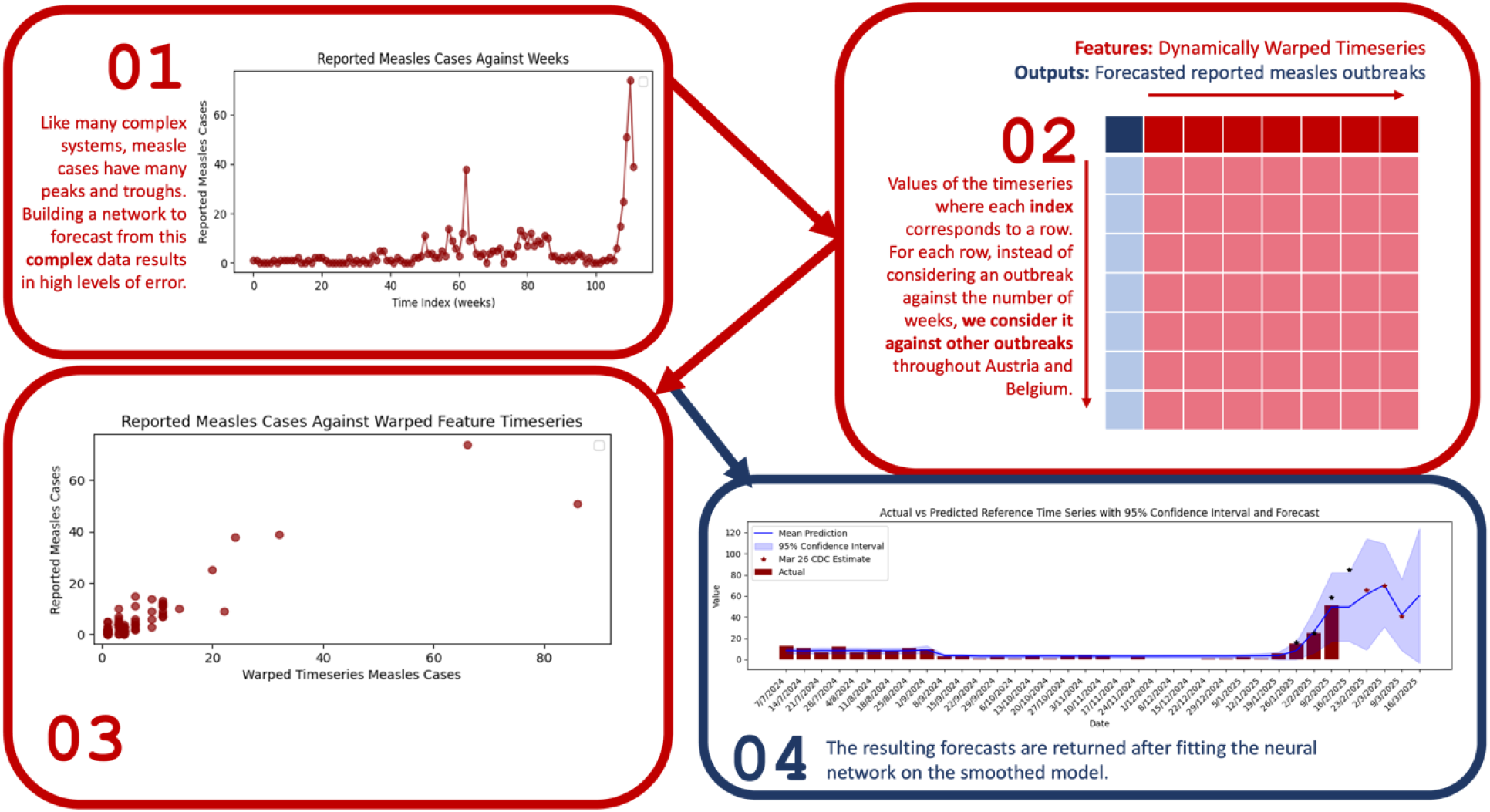
Dynamic time warping (DTW) of the feature timeseries results in a more simplistic model for learning.

### Feedforward neural network (FNN)

An FNN was chosen as the foundation of our model with feature inputs taken as the set of twelve historical outbreak timeseries and a one-week delayed timeseries of (past) reported weekly measles cases across the USA. The target output is taken as the number of weekly reported measles cases across the USA (Figure 2a). The model used two linear hidden layers (dim 64, dim 32) equipped with the ReLU activation function. In line with standard optimisation for the FNN, loss was defined as the mean-squared error of the FNN prediction, *FNN*(*x*(*t*_*i*_)), against the true weekly reported measles cases in the USA, *y*(*t*_*i*_).

**Figure 2:**
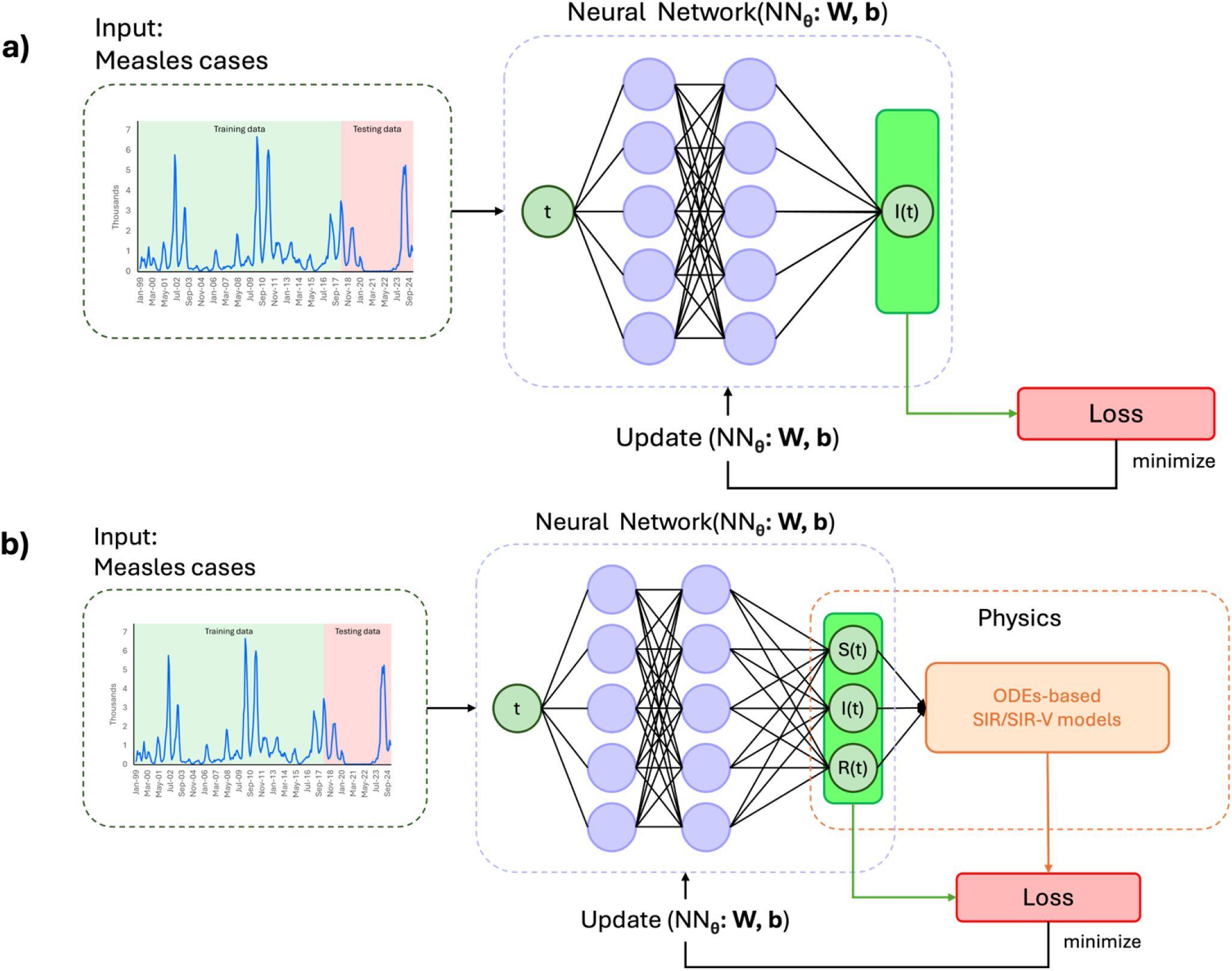
Architecture for a) our simple, feed-forward neural network taking historical timeseries of measles cases from Austria and Belgium as features into the network and b) our proposed novel neural network incorporating dynamics of SIR models into the latent space of the network.

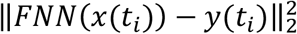

Predictions by the FNN were updated through an iterative process of choosing weights on combinations of the ReLU functions in the hidden layers of the network such that the loss was minimised.

### Biologically informed neural network (BINN)

We now introduce a new network architecture we call a BINN, constructed on the premise of a Physics Informed Neural Network but using mathematical models describing biological dynamics in the latent space of the network (Figure 2b). For this application, a coupled system of differential equations describing an SIR model was incorporated into the latent space of the neural network. This was achieved by redefining the loss function governing both the quantity of measles infections at time index *i* and the change in the number of measles cases,

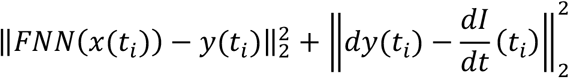

where the value *dy*(*t*_*i*_) is calculated from the actual change in measles cases at our current time step in the timeseries and 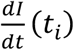 is the expected value on the change of the number of active cases at time *t*_*i*_, informed by the coupled system of ordinary differential equations describing an SIR model, namely,

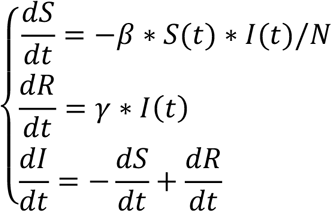

### Model testing and training

We split our target timeseries (weekly reported measles cases in the U.S.) into a 70% training set and a 30% test set. Unlike common classification problems, the test set here is chosen as the last 30% of the timeseries of reported measles cases to preserve chronology. Optimisation was performed via gradient descent using the built-in python optimiser ADAM with learning rate 0.001. Convergence for the training set was observed within 100 epochs. Mean model performance was evaluated using a bootstrap approach which sampled, with replacement, 100 snips of the feature time series the same length as the test set. Confidence intervals were also calculated through the bootstrap method.

### Code availability

All analysis was performed using Python and all code is available on UQ eSpace.

## RESULTS

### Measles outbreaks in the USA, 2025

2025 represents a significant outbreak year in the history of measles in the USA (Figure 3). This was largely driven by an outbreak originating from Gaines County, West Texas at the start of 2025. Gaines county represents a pocket of Texas with lower that the stipulated 95% coverage rate (82%) for the measles mumps rubella (MMR) vaccine (Figure 4). Since commencing, the outbreak has now spread to other counties within Texas as well as other states.

**Figure 3:**
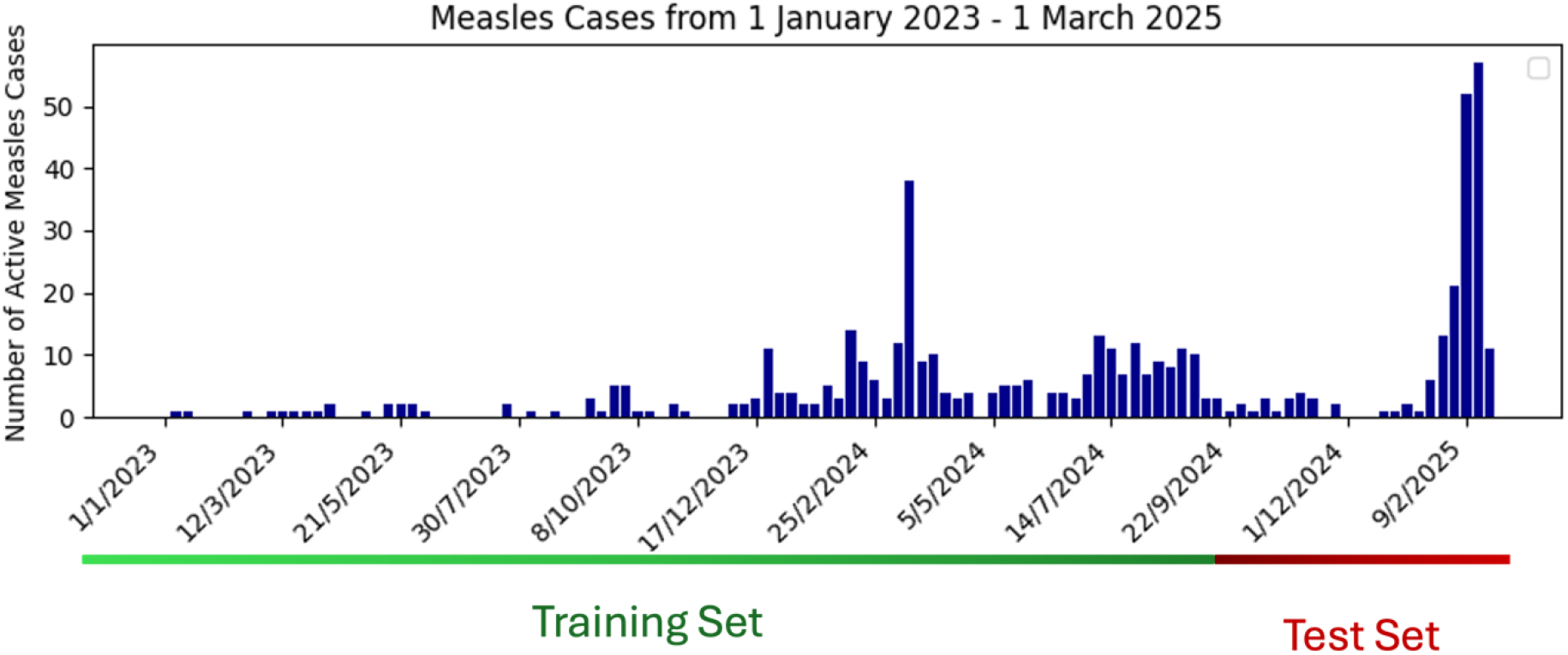
Yearly measles cases recorded in the USA from 2000-2025. Data is derived from the CDC accessed on 6 March 2025.

**Figure 4:**
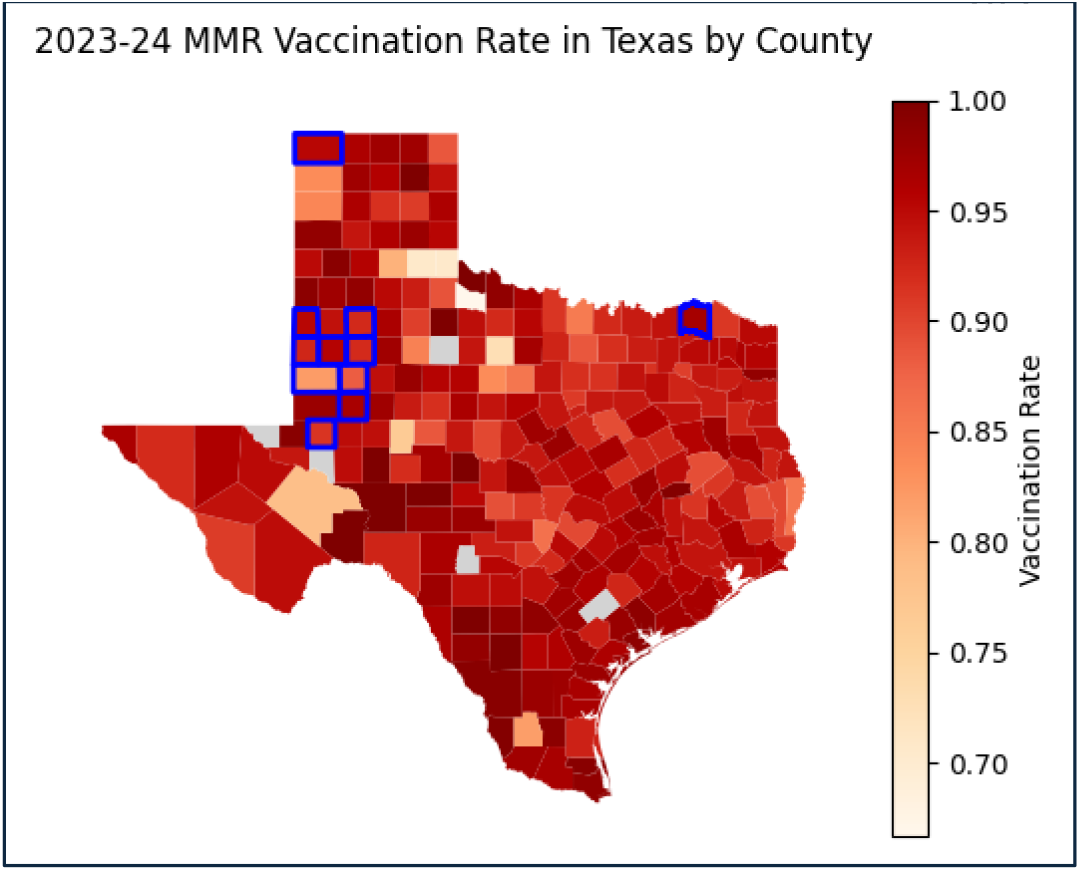
Measles vaccination rates for kindergarteners by county results of the 2023-2024 Texas Annual Report of Immunization. Gaines County reports an 82% vaccination rate compared to the Texas state vaccination rate at 94%. Blue outlines indicate counties with measles cases traced back to the Gaines County measles outbreak.

### An FNN to model US measles cases in 2025

To model measles cases in the USA in 2025 an FFN was created using historical measles outbreak data from Europe for feature selection and a one-week time delayed weekly measles outbreak timeseries from Texas. Weeks from 1 January 2023 to 7 July 2024 were used as training data whilst weeks from 7 July 2024 through to 1 March 2025 correspond to the test set. Figure 5a shows that across the 34 weeks of test data measles cases were predicted with a mean-squared error of less than 2 (1.1060) cases. To investigate the prediction horizon, we generated a forecast 5 weeks using the same bootstrap method as was performed on the test set (Figure 5a). The model was trained and tested on weeks through the 1 March 2024 and thus anything after this date represents a forward prediction. Furthermore, data updated after 6 March 2025 from the CDC were not used by the model to make predictions. Figure 5a shows that in the instance of forward prediction the model is also highly accurate.

**Figure 5:**
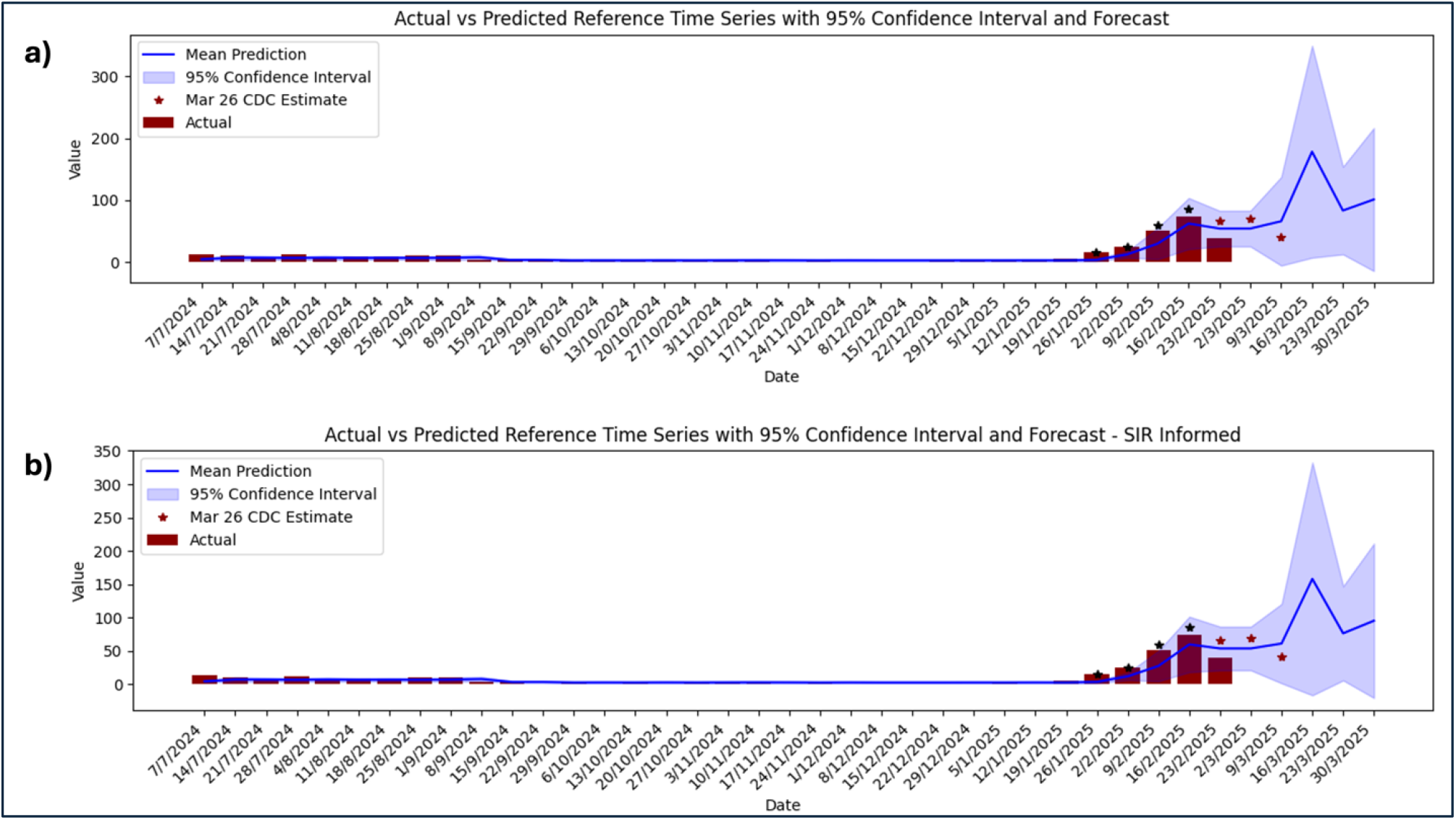
Actual vs. Predicted number of USA measles cases from 7/7/2024 to 30/3/2025 (test set) using the a) FNN and b) BINN with counts estimated as of 6 March 2025. The 95% confidence interval is shown in pale blue, and the forecast is shown in bright blue. Current CDC estimates of weekly measles cases that were obtained after model development on 22 March 2025 are shown as red stars.

### A Biologically Informed Neural Network (BINN) to model US measles cases in 2025

Using the same training and test set as the FNN, the BINN architecture was employed with an SIR model governing the expected change in measles cases at each time step. Figure 5b shows that across the training data measles cases were predicted with a mean-squared error across the 34 weeks represented in the test set was 1.1451. Compared to FNN forecasts where a second peak of similar height is expected, the BINN forecasts nearly identical values to the FNN. In sum, these data indicate that the BINN did not improve the prediction of 2025 measles cases in the USA beyond that already provided by the FNN.

## DISCUSSION

The 2025 resurgence of measles in the USA, a disease previously declared eliminated, is the direct result of declining vaccination rates. The current Texas outbreak has resulted in more than 300 infections and, tragically, two deaths. Modelling the current outbreaks of measles in the USA in real-time is essential for organizing vaccination campaigns and allocating resources effectively. Here, we show that a simple FNN, trained on historical data, can accurately predict 2025 measles cases in the USA. Indeed, in addition to its accurate performance on the test set, our model maintains a reasonable 2-week forecasting accuracy without updates on the feature data as we continue to observe reported measles cases live from the CDC.

Essential to the success of this model was using dynamic time warp to address problems of time series matching. Dynamic time warping works by finding the most optimal alignment of each historical timeseries, or path of points, to the reference timeseries by calculating the total distance between the reference and historical timeseries for every possible alignment in the space. Accordingly, this technique allowed us to use historical outbreak timeseries, which were measured on the order of one month and with variable outbreak length, for feature selection. Incorporation of dynamic time warping (DTW) into simple neural networks has been explored in previous studies involving classification of similar images with varying orientations where images were converted to signals prior to being processed in the DTW layer ^14^. In line with our conclusions, the authors find that a simple DTW FNN outperformed current literature standards using Recurrent Neural Networks (RNN) (e.g. Long-Short Term Memory). We theorise that the reason an RNN performs poorly in this context is that it assumes the timeseries will follow very recent trends, which is not true for highly contagious outbreaks (e.g. measles) resulting in quick peaks of cases ^15^. To the best of our knowledge dynamic time warping has not previously been used in prior models developed to predict measles outbreaks. Dynamic time warping thus represents a powerful, and potentially underutilised, tool in infectious disease forecasting.

The use of dynamic time wrapping herein enabled us to use historical data as a feature to develop a neural network. Prior work in this field has focussed on using features like vaccination rates, socioeconomic status and other similar factors for machine learning models that predict measles outbreaks ^7-9^. However, such an approach is not without its limitations. Socioeconomic data, for instance, may not be regularly updated or uniformly available across regions, making it challenging to maintain accuracy and relevance. Additionally, collection, cleaning, preprocessing and integrating diverse data types requires significant time and computational resources. Together, this complexity could slow down model development and limit its relevance for real-time applications. The use of scaled historical data circumvents many of these challenges and offers readily available data for feature selection and training. This is particularly true if historical data is drawn from the same or equivalent countries (in terms of socioeconomic status, vaccination rates etc.) as was the case herein. However, even with the use of dynamic time warping, historical data may not be suitable to use for feature selection in all viral contexts. For example, one could imagine that the use of outbreak data of SARS-CoV-2 in 2020 would provide poor data for feature selection for a model designed to predict outbreaks of SARS-CoV-2 in 2025. Not only has SARS-CoV-2 mutated over time (altering transmission dynamics) but population immunity, public health policy and societal behaviour is now (in 2025) dramatically different to that in 2020. Thus, whilst the virus, public health policy and societal behaviour have remained relatively consistent in the context of measles ^1^, these remain important caveats to the accurate use of historical training data for other more variable outbreak contexts.

Neural networks are historically difficult to adopt into public health policy as they represent a ‘black box’ approach and thus may not provide the necessary level of transparency required to inform public policy. Here, we use the most simplistic architecture in deep learning, the hidden layer feed forward neural network, as our model foundation. We further seek to add transparency to our models by incorporating known population dynamics of measles outbreaks studied through the SIR framework ^16-19^. Interestingly, the incorporation of these SIR model principles into the neural network created herein did not result in increased accuracy. SIR models are a well-established and often highly accurate approach to modelling outbreaks of infectious disease. We speculate that, for longer training, SIR model principles do not improve the performance of the FNN because these principles are already incorporated in the data used for feature selection. That is, by using real-world historical data (the dynamics of which are already governed by the principles of SIR) the incorporation of SIR principles in our model is redundant. If true, this represents an additional advantage of using historical data for feature selection when modelling infectious disease. On the other hand, given that accuracy was not reduced in the BINN, our results also demonstrate the potential benefit of SIR drivers in deep networks where historical data is scarce or missing.

The models developed herein suggest that without a dramatic change in public health policy the number of measles cases in the USA will continue to grow. This puts all unvaccinated individuals at risk, including those who are too young to receive the measles vaccine. We thus need strong promotion and advocacy of the measles mumps rubella (MMR) vaccine at all levels of society. Indeed, there are lessons to be learned from past measles outbreaks in Orthodox Jewish communities in New York. Here, an essential part of the outbreak response was the employment of nurses as ambassadors for immunization, bringing evidence-based health information to the Orthodox Jewish community in a culturally sensitive manner ^20^. In sum, our data suggest that a strong public health response is needed now more than ever to prevent the short- and long-term consequences of a measles infection and advert any more unnecessary loss of life.

## Data Availability

Data and code will be made available upon request after publication of the manuscript. Links to data access are available on UQ eSpace.

